# Immune response of primary and booster immunity of SARS-CoV-2 vaccination among patients with chronic liver disease

**DOI:** 10.1101/2022.11.12.22282242

**Authors:** Ruixin Song, Chao Yang, Jiayin Wang, Qianqian Li, Jing Chen, Kai Sun, Hongmin Lv, Yankai Yang, Jing Liang, Qing Ye, YanYing Gao, Jun Li, Ying Li, Junqing Yan, Ying Liu, Tao Wang, Changen Liu, Fei Wang, Weili Yin, Huiling Xiang

## Abstract

**Aim:** we examined the humoral immune response and antibody dynamics after primary and booster vaccination against severe acute respiratory syndrome coronavirus 2 (SARS-CoV-2) among patients with chronic liver disease (CLD).

**Methods:** We enrolled patients with confirmed CLD and SARS-CoV-2 vaccination primary or booster immunity. Following primary or booster immunity, serological samples were gathered to detect novel coronavirus neutralizing antibodies (nCoV NTAb) and novel coronavirus spike receptor-binding proteins (nCoV S-RBD).

**Results:** The positive rate of nCoV NTAb was 60.1% in Primary and 87.6% in Booster (P<0.001). The median level of nCoV NTAb was 11.6 AU/mL in Primary and 31.6 AU/mL in Booster (P<0.001). The positive rate of nCoV S-RBD was 70.0% in Primary and 91.2% in Booster (P<0.001). The median level of nCoV S-RBD was 21.7 AU/mL in Primary and 110.6 AU/mL in Booster (P<0.001). Compared to the antibody level of primary immunity, 21 patients in whom SARS-CoV-2 vaccine antibodies were detected after both primary and booster immunizations showed an increase of 4.4 and 5.9 times in nCoV NTAb and nCoV S-RBD, respectively.

**Conclusion:** Patients with CLD show improved humoral immune response after completing primary and booster immunity of SARS-CoV-2 vaccines, while booster immunity further improves the positive rate and antibody level of patients with CLD.

---

As of April 2023, 762 million people have been proved to infect Coronavirus Disease 2019 (COVID-19), and 6.9 million people have died all over the world[1]. Vaccines of severe acute respiratory syndrome coronavirus 2 (SARS-CoV-2) has been implemented successively. In the 20-59-year-old age group, BNT162b2 and CoronaVac vaccination avoided mortality after COVID-19 diagnosis by 91.7%[2]. Those with chronic liver disease (CLD) have congenital and adaptive immune cell suppression[3], a higher risk of infection and mortality[4, 5]; hence, SARS-CoV-2 vaccination should be targeted at these patients. After completing the primary immunization for participants with CLD, the positive rate of novel coronavirus neutralizing antibody (nCoV NTAb) and novel coronavirus spike receptor-binding domain antibody (nCoV S-RBD) was 87.8% and 87.8%[6], respectively. Considering the diminishing effectiveness of vaccines[7, 8], the World Health Organization (WHO) recommended a booster dose vaccination. Compared to primary immunization of SARS-CoV-2 vaccines, the effectiveness of a booster dose in preventing the occurrence of COVID-19 is 95.3%[9]. Notably, three doses reduce the hospitalization rate, disease severity, and mortality[10]. Several studies have discussed the humoral immune response of patients with CLD after primary immunity with SARS-CoV-2 vaccines. However, data regarding booster immunity on the humoral immune response in CLD are yet lacking. This study investigated the humoral immunity and antibody dynamics with SARS-CoV-2 vaccines among CLD after completing primary and booster immunity. Hence, this study contributes to advancing vaccination tactic about SARS-CoV-2.

## 1. Methods

### 1.1 Inclusion and exclusion criteria

This study enrolled patients visiting the hepatology outpatient clinic of the Third Central Hospital of Tianjin in China between January 2021 and June 2022. Here are the criteria for inclusion: 1. Age ≥18 years; 2. CLD diagnosed by clinical data or medical history; 3. Completion of primary immunity or booster immunity of SARS-CoV-2 vaccines; 4. Consent to take part in the research and comply with research procedures.

It was excluded from this study if you were pregnant, lactating, active or previous COVID-19 infection, previously undergone liver transplantation, immunosuppressive or had an immunodeficiency disorder [including human immunodeficiency virus (HIV) infection], the use of systemic immunosuppressive medications, systemic immunoglobulins or immune boosters within the last three months.

### 1.2 Diagnostic basis of CLD

The definition of CLD is liver disease that lasts more than six months and included chronic liver inflammation, irrespective of cirrhosis. Cirrhosis diagnostic criteria are based on the Cirrhosis Diagnosis and Treatment Guidelines[11]

### 1.3 Vaccine types and injection protocol

The types of vaccines in this study included BBIBP-CorV vaccine (Beijing Institute of Biological Products Co., Ltd, China), CoronaVac vaccine (Beijing Sinovac Life Sciences Co., Ltd, China), CanSinoBio Ad5-nCoV vaccine (CanSino Biologics Inc. China), and Zifivax vaccine (CHO cells) (Anhui Zhifei Longcom Biopharmaceutical Co., Ltd, China). The immunization protocol was based on the SARS-CoV-2 immunization Guidelines[12].

CoronaVac and BBIBP-CorV vaccines are inactivated and require two doses at an interval of 3–8 weeks, with a booster dose at least 6 months apart. CanSinoBio Ad5-nCoV vaccine is an adenovirus vector vaccine that requires one dose; a booster dose is a minimum of six months except the primary dose. The Zifivax vaccine is a recombinant subunit vaccine requiring three doses, 4–8 weeks intervals between each dose, and does not require a booster dose.

### 1.4 Clinical data collection

Clinical data included baseline demographics [age, gender, race, and body mass index (BMI)], liver disease-related situation (etiology, absence or presence cirrhosis, and Child-Pugh class), comorbidity-related situation (hypertension, diabetes, coronary artery disease, asthma, and arrhythmia), SARS-CoV-2 vaccination information (the type of vaccine and the date of completion primary or booster immunity), laboratory examination-related indicators (alanine aminotransferase, aspartate aminotransferase, albumin, gamma-glutamyl transpeptidase, and alkaline phosphatase).

### 1.5 Assessment of immune response

Serological specimens were collected after completion of primary or booster immunity of SARS-CoV-2 vaccines to detect nCoV NTAb and nCoV S-RBD [competitive combination chemiluminescence immunoassay (CLIA) method by Shenzhen Mindray Bio-Medical Electronics Co., Ltd, China]. Based on the instructions provided by the manufacturer, antibody level over 10.0 AU/mL was thought positive, while antibody level less than 10.0 AU/mL was thought negative.

### 1.6 Safety assessment

Local and systemic side effects of patients with CLD following primary and booster immunization of SARS-CoV-2 vaccinations were reported. The local side effects at the injection site comprised ache, swelling, redness, induration, pruritus at the injection site, and their severity. Systemic side effects comprised fever, dizziness, fatigue, nausea, vomiting, decreased appetite, myalgia, arthralgia, oropharyngeal pain, cough, allergy, dyspnea, syncope, pruritus (non-injection site), and their severity.

### 1.7 Statistical analysis

Numerical variables were portrayed by mean (standard deviation) or median (IQR) depending on whether they conformed normal distribution. The frequencies and percentages of each variables were used to represent the categorical variables. Numerical variables were assessed by t-test and nonparametric test to evaluate the Statistical difference. Categorical variables were evaluated for statistically significant differences between groups using Pearson’s χ^2^ test and Fisher’s exact test. P-values below 0.05 were thoughted the statistical difference when using the two-sided test. SPSS 24.0 was applied to the statistical analysis. The images were generated using Origin 2023.

### 1.8 Ethical Statements

Prior to registration, each participant provided written informed consent. The research is approved by the Ethics Committee of Tianjin Third Central Hospital in China. This research protocol adheres to the ethical guidelines outlined in the 1975 Declaration of Helsinki (Approval number:IRB 2021-027-01).

## 2. Results

### 2.1 Baseline demographic and clinical characteristics

316 patients completed SARS-CoV-2 vaccines and were divided into primary and booster immunity groups according to the status of vaccination. In this study, the primary immunity group was referred to as Primary, and the booster immunity group was referred to as Booster. Primary included 203 patients who completed the primary immunity of SARS-CoV-2 vaccination, and Booster included 113 patients who completed booster immunity of SARS-CoV-2 vaccination (Figure 1). The median age of the primary and booster groups was 54.0 (IQR: 42.0-62.0) years and 52.0 (IQR: 42.5-59.5) years (P=0.446). Males accounted for 56.2% and 62.8% in Primary and Booster (P=0.284). Regarding etiology, CLD was caused by hepatitis B virus, and the proportion of hepatitis B was 67.0% and 86.7%, respectively (P=0.001). The median time between primary immunity completion to serological specimen collection was 55 (IQR: 32-102) days, and the median time between booster immunity completion and serological specimen collection was 102 (IQR: 45-145) days (P<0.001) (Table 1).

**Figure 1.**
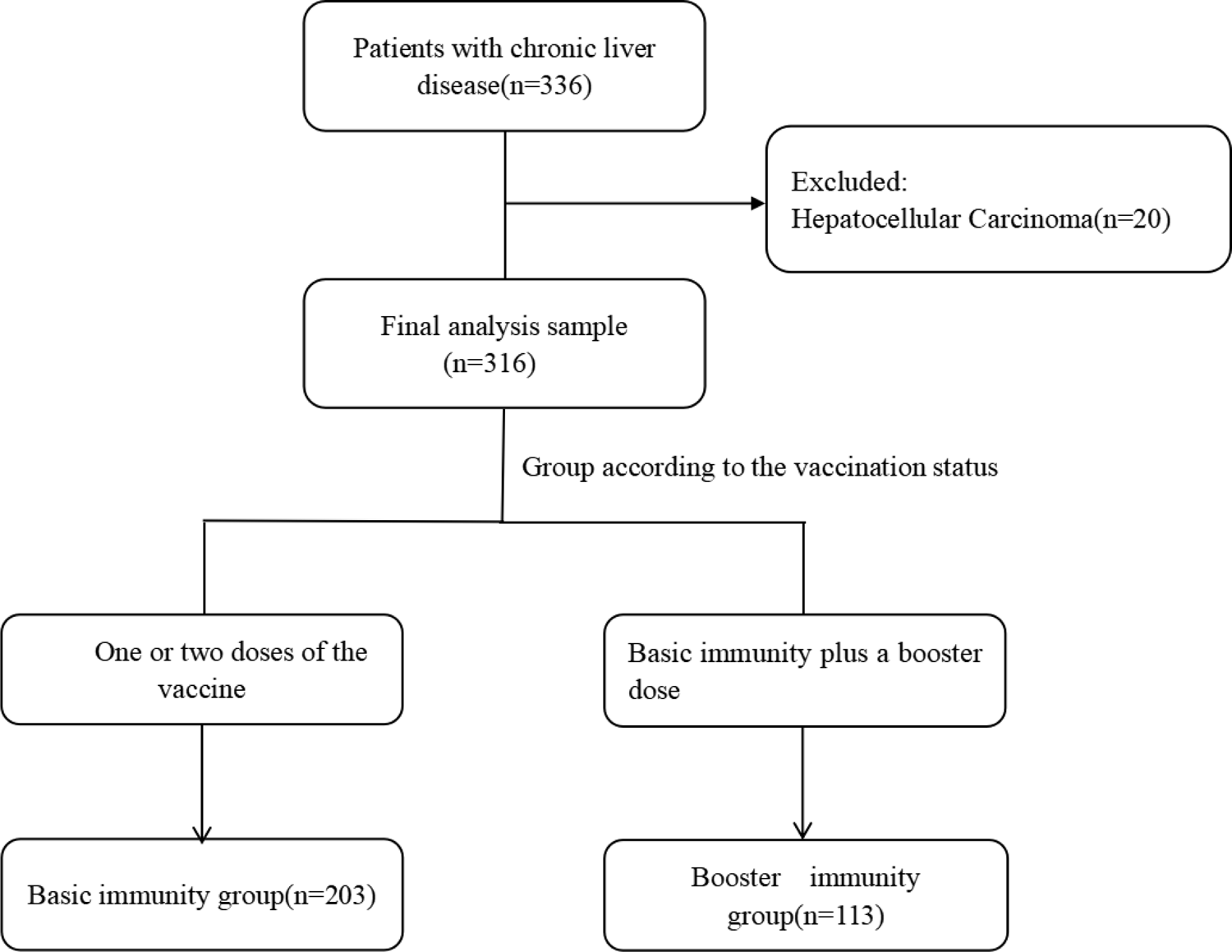
Study flow chart.

**Table 1.** Baseline demographic and clinical characteristics

| **Characteristic** | **Primary** | **Booster** | **P-Value** |
| --- | --- | --- | --- |
| **N** | 203 | 113 |  |
| **Age,year** | 54.0(42.0-62.0) | 52.0(42.5-59.5) | 0.446 |
| **Gender,male** | 114(56.2) | 71(62.8) | 0.284 |
| **Race , Han** | 199(98.0) | 110(97.3) | 0.608 |
| **BMI** | 24.7±3.5 | 24.7±3.5 | 0.960 |
| **Etiology** |  |  | 0.001 |
| **HBV** | 136(67.0) | 98(86.7) |  |
| **HCV** | 19(9.4) | 4(3.5) |  |
| **ALD** | 10(4.9) | 1(0.9) |  |
| **AIH、 PBC** | 10(4.9) | 6(5.3) |  |
| **others** | 28(13.8) | 4(3.5) |  |
| **Presence of cirrhosis** | 96(47.3) | 51(45.1) | 0.726 |
| **Child-Pugh class** |  |  | 0.848 |
| **A** | 91(94.8) | 48(94.1) |  |
| **B** | 2(2.1) | 2(3.9) |  |
| **C** | 3(3.1) | 1(2.0) |  |
| **Comorbidities** | 78(38.4) | 38(33.6) | 0.465 |
| **Hypertension** | 48(23.6) | 20(17.7) | 0.254 |
| **Diabetes** | 30(14.8) | 12(10.6) | 0.308 |
| **CAD** | 4(2.0) | 5(4.4) | 0.290 |
| **Asthma** | 4(2.0) | 0(0.0) | 0.184 |
| **Arrhythmia** | 1(0.5) | 1(0.9) | 0.680 |
| **Vaccine category** |  |  | 0.004 |
| **BBIBP-CorV** | 96(47.3) | 45(39.8) |  |
| **CoronaVac** | 92(45.3) | 51(45.1) |  |
| **CansinoBio** | 15(7.4) | 11(9.7) |  |
| **CHO cell Vac** | 0(0.0) | 6(5.3) |  |
| **Time interval** | 55(32-102) | 102(45-145) | < 0.001 |
| **Liver function** |  |  |  |
| **ALT(U/L)** | 24.0(17.8-33.0) | 22.5(16.0-36.0) | 0.327 |
| **AST(U/L )** | 23.0(19.0-29.0) | 22.5(18.0-31.0) | 0.637 |
| **ALB ( g/L)** | 47.8(45.6-49.7) | 48.0(46.1-51.6) | 0.293 |
| **GGT(U/L)** | 25.0(17.0-46.0) | 30.5(17.8-49.5) | 0.407 |
| **ALP(U/L)** | 76.0(63.0-94.0) | 30.5(17.8-49.5) | 0.917 |

Data was presented as median (interquartile range), Mean±standard deviation, or frequencies(percentage). Vaccine category referred to the type of first dose COVID-19 vaccine. Time interval meant the time (days) between the completion of primary or booster immunity and the collection of serological samples. BMI, Body Mass Index; HBV, Hepatitis B virus; HCV, Hepatitis C virus; ALD, Alcoholic liver disease; AIH, Autoimmune hepatitis; PBC, Primary biliary cholangitis; CAD, Coronary artery disease; ALT, Alanine aminotransferase; AST, Aspartate aminotransferase; ALB, Albumin; GGT, Gamma-glutamyl transpeptidase; ALP, Alkaline phosphatase.

### 2.2 Immune response assessment

#### 2.2.1 nCoV NTAb

The positive rate of nCoV NTAb was 60.1% in Primary and 87.6% in Booster (P<0.001) (Table 2 and Figure 2A). A booster dose increased nCoV NTAb positivity rate by 27.5%. The median level of nCoV NTAb was 11.6 (IQR: 8.1-20.3) AU/mL in Primary and 31.6 (IQR: 15.4-97.0) AU/mL in Booster (P<0.001) (Table 2 and Figure 2B). A booster dose increased the level of nCoV NTAb by 2.7-fold.

**Figure 2.**
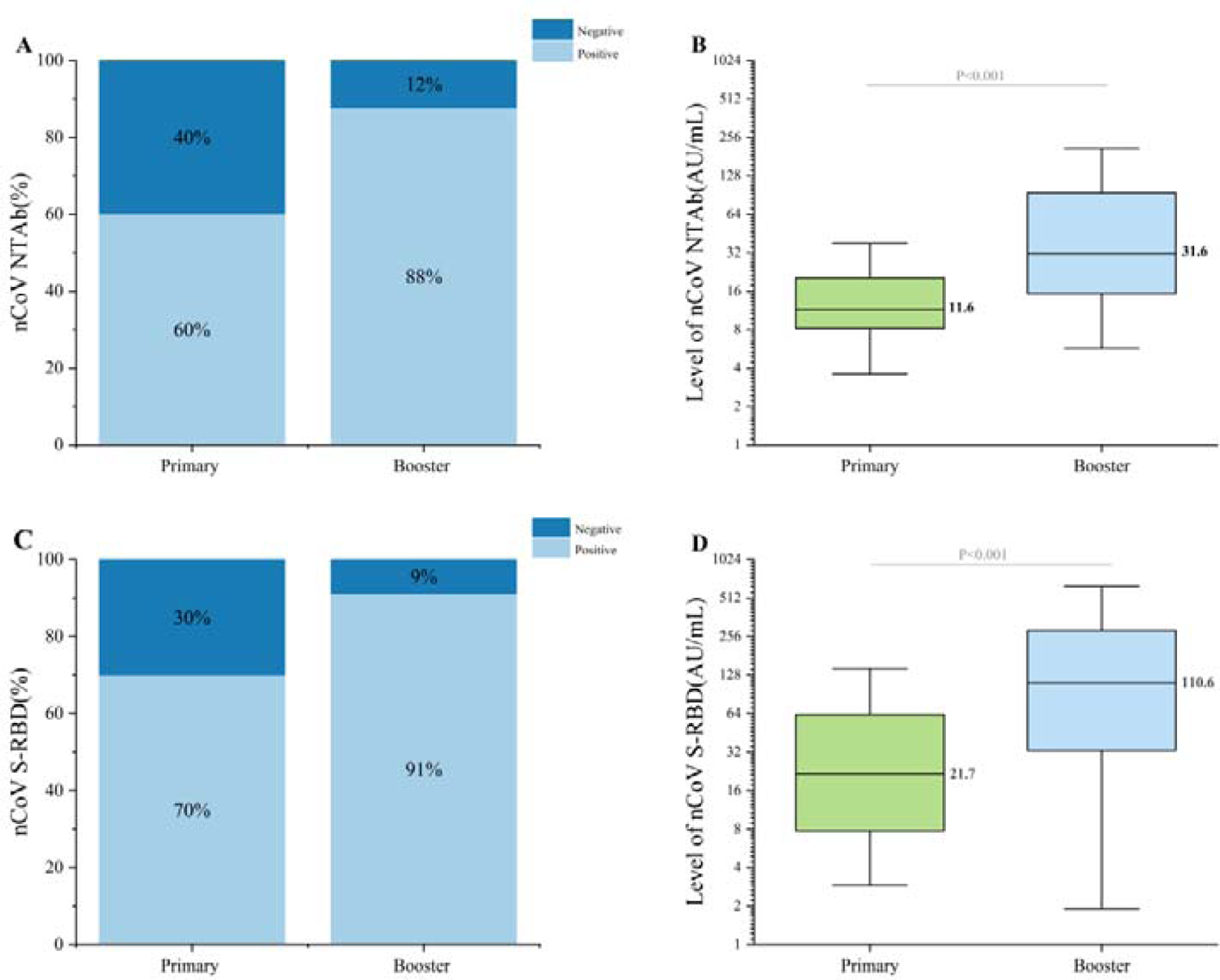
The humoral immune response of COVID-19 vaccine in Primary and Booster groups among patients with chronic liver disease. Figure A revealed the positive rate of nCoV NTAb in Primary and Booster groups. Figure B revealed the level of nCoV NTAb in Primary and Booster groups. Figure C revealed the positive rate of nCoV S-RBD in Primary and Booster groups. Figure D revealed the level of nCoV S-RBD in Primary and Booster groups. Antibody level over 10.0 AU/mL was thought positive, while antibody level less than 10.0 AU/mL was thought negative. nCoV NTAb, novel coronavirus neutralizing antibody; nCoV S-RBD, novel coronavirus spike receptor-binding domain antibody.

**Table 2.** The humoral immune response of COVID-19 vaccine in Primary and Booster groups among patients with chronic liver disease.

|  | **Primary** | **Booster** | **P-Value** |
| --- | --- | --- | --- |
| **nCoV NTab** |  |  |  |
| The positive rate | 122(60.1) | 99(87.6) | <0.001 |
| Antibody level | 11.6(8.1-20.3) | 31.6(15.4-97.0) | <0.001 |
| **nCoV S-RBD** |  |  |  |
| The positive rate | 142(70.0) | 103(91.2) | <0.001 |
| Antibody level | 21.7(7.7-63.0) | 110.6(31.0-285.4) | <0.001 |

Data was presented as median (interquartile range) or frequencies(percentage). Antibody level above 10.0 AU/mL was regarded as positive, while antibody level below 10.0 AU/mL was regarded as negative. nCoV NTab, novel coronavirus neutralizing antibody; nCoV S-RBD, novel coronavirus spike receptor-binding domain antibody.

Subgroup analysis: The humoral immune response of non-cirrhosis and cirrhosis was compared in Primary and Booster. After Primary, the positive rate of nCoV NTAb was 58.9% and 61.5% in non-cirrhosis and cirrhosis patients (P=0.775), respectively, and the median level was 11.3 (IQR: 7.8-20.3) AU/mL and 12.1 (IQR: 8.6-20.5) AU/mL in non-cirrhosis and cirrhosis (P=0.577), respectively. After Booster, the positive rate of nCoV NTAb was 93.5% and 80.4% in non-cirrhosis and cirrhosis (P=0.045), respectively, and the median level was 31.4 (IQR: 19.5-96.0) AU/mL and 32.3 (IQR: 11.3-99.7) AU/mL in non-cirrhosis and cirrhosis, respectively (P=0.441) (Table 3 and Figure 3A).

**Figure 3.**
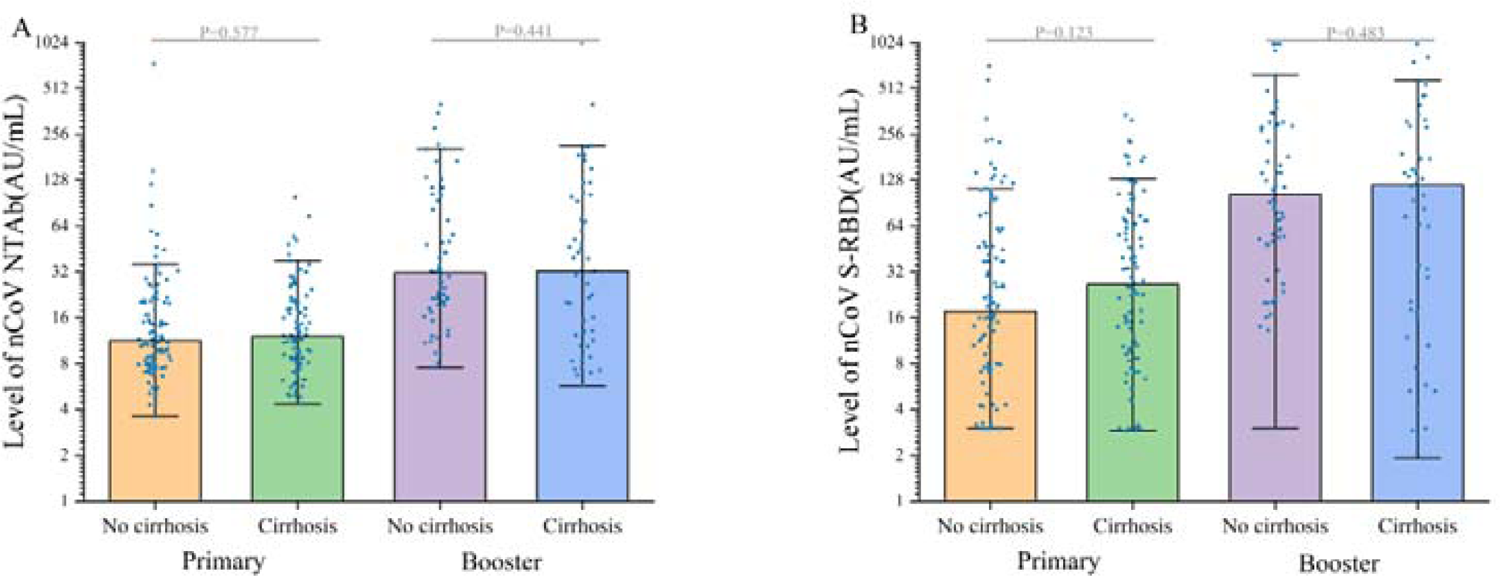
The level of nCoV NTAb and nCoV S-RBD in Primary and Booster groups among patients with no cirrhosis and cirrhosis. Figure A showed the level of nCoV NTAb and Figure B showed the level of nCoV S-RBD. nCoV NTAb, novel coronavirus neutralizing antibody; nCoV S-RBD, novel coronavirus spike receptor-binding domain antibody.

**Table 3.**
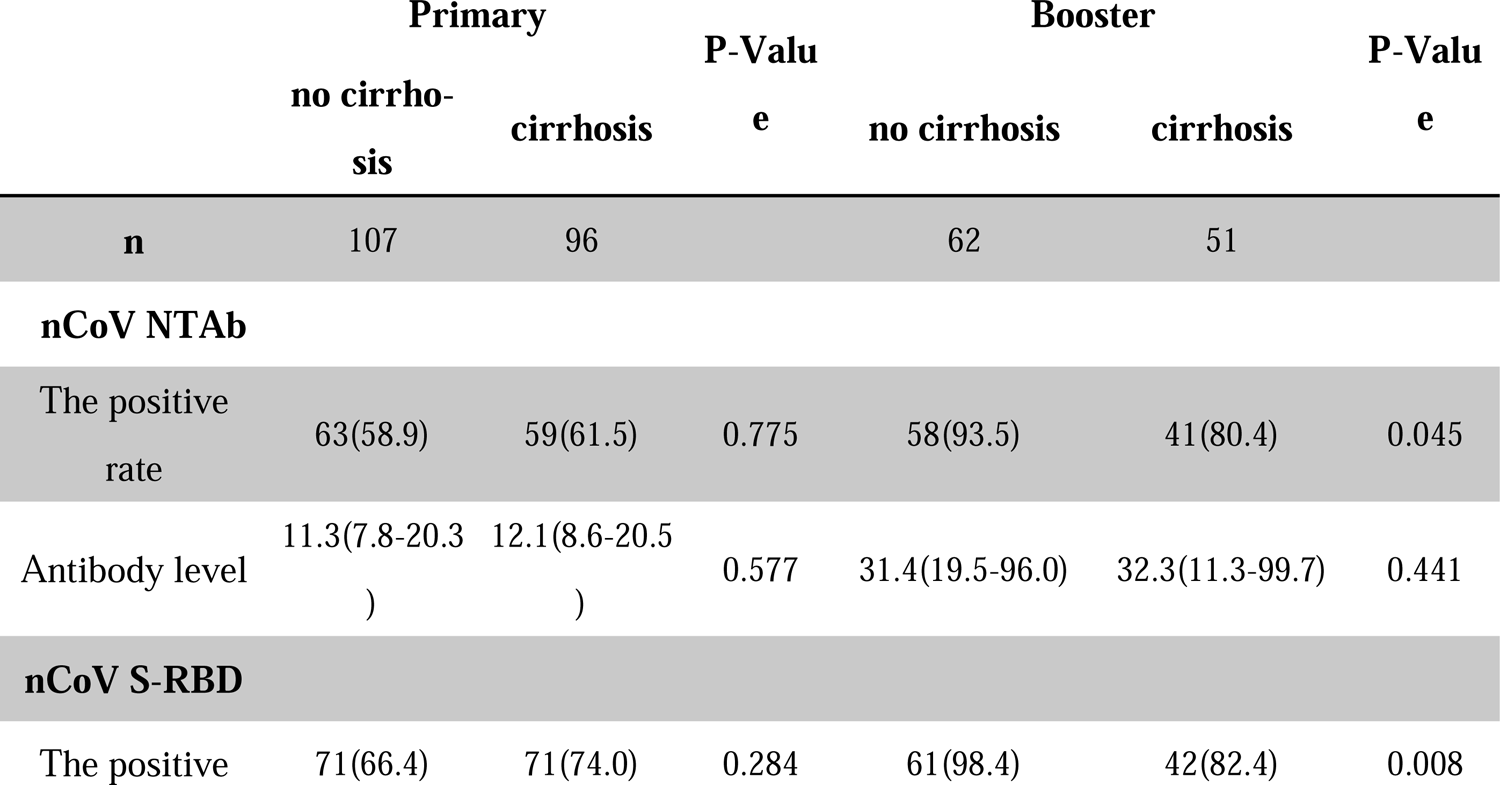
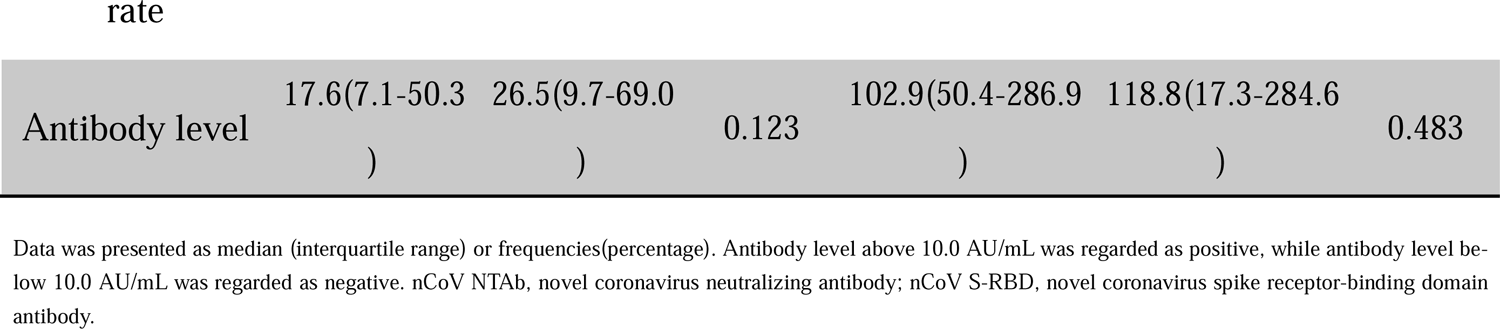
The humoral immune response of COVID-19 vaccine in Primary and Booster groups among no cirrhosis and cirrhosis patients.

|  | Primary |  | P-Value | Booster |  | P-Value |
| --- | --- | --- | --- | --- | --- | --- |
|  | no cirrhosis | cirrhosis |  | no cirrhosis | cirrhosis |  |
| n | 107 | 96 |  | 62 | 51 |  |
| **nCoV NTab** |  |  |  |  |  |  |
| The positive rate | 63(58.9) | 59(61.5) | 0.775 | 58(93.5) | 41(80.4) | 0.045 |
| Antibody level | 11.3(7.8-20.3) | 12.1(8.6-20.5) | 0.577 | 31.4(19.5-96.0) | 32.3(11.3-99.7) | 0.441 |
| **nCoV S-RBD** |  |  |  |  |  |  |
| The positive | 71(66.4) | 71(74.0) | 0.284 | 61(98.4) | 42(82.4) | 0.008 |
| Antibody level | 17.6(7.1-50.3<br>) | 26.5(9.7-69.0<br>) | 0.123 | 102.9(50.4-286.9<br>) | 118.8(17.3-284.6<br>) | 0.483 |

Data was presented as median (interquartile range) or frequencies(percentage). Antibody level above 10.0 AU/mL was regarded as positive, while antibody level below 10.0 AU/mL was regarded as negative. nCoV NTAAb, novel coronavirus neutralizing antibody; nCoV S-RBD, novel coronavirus spike receptor-binding domain antibody.

#### 2.2.2 nCoV S-RBD

The positive rate of nCoV S-RBD was 70.0% in Primary and 91.2% in Booster (P<0.001) (Table 2 and Figure 2C). A booster dose increased nCoV S-RBD positivity rate by 21.2%. The median level of nCoV S-RBD was 21.7 (IQR: 7.7-63.0) AU/mL in Primary and 110.6 (IQR: 31.0-285.4) AU/mL in Booster (P<0.001) (Table 2 and Figure 2D). A booster dose increased the level of nCoV S-RBD by 5.1 times.

Subgroup analysis: After Primary, the positive rate of nCoV S-RBD was 66.4% and 74.0% (P=0.284), and the median level was 17.6 (IQR: 7.1-50.3) AU/mL and 26.5 (IQR: 9.7-69.0) AU/mL in non-cirrhosis and cirrhosis, respectively (P=0.123). After Booster, the positive rate of nCoV S-RBD was 98.4% and 82.4% (P=0.008), and the median level was 102.9 (IQR: 50.4-286.9) AU/mL and 118.8 (IQR: 17.3-284.6) AU/mL in non-cirrhosis and cirrhosis, respectively (P=0.483) (Table 3 and Figure 3B).

#### 2.2.3 Antibody dynamics after booster immunity

A total of 21 patients underwent antibody detection of SARS-CoV-2 vaccines after both primary and booster immunity to further confirm the dynamic changes in antibodies after booster immunity. Subsequently, the positive rate of nCoV NTAb increased from 57.1% to 85.7%, and that of nCoV S-RBD increased from 66.7% to 90.5% (Table 4 and Figure 4A). The median level of nCoV NTAb increased from 11.1 (IQR: 8.4-16.0) AU/mL to 48.8 (IQR: 16.8-143.4) AU/mL after booster immunity. The median level of nCoV S-RBD was increased from 24.3 (IQR: 7.2-48.9) AU/mL to 142.5 (IQR: 55.8-424.9) AU/mL (Table 4 and Figure 4B). Compared to the antibody level of primary immunity, the median level of nCoV NTAb and nCoV S-RBD was increased by 4.4 and 5.9 times, respectively. In conclusion, after booster immunity of SARS-CoV-2 vaccine, the positive rate and antibody level in patients with CLD can be further improved based on primary immunity. Among 21 patients, 9 were negative for nCoV NTAb after primary immunity, while 6 cases were transformed positive after booster immunity, and the positive conversion rate of nCoV NTAb was 66.7% (Figure 4C). Furthermore, 7 patients were negative for nCoV S-RBD after primary immunity, while 5 were transformed into positive after booster immunity, and the positive conversion rate was 71.4% (Figure 4D). The positive conversion rate among patients with primary immunity failure was also improved after booster immunity, such that patients with primary immunity failure can acquire immunity to COVID-19.

**Figure 4.**
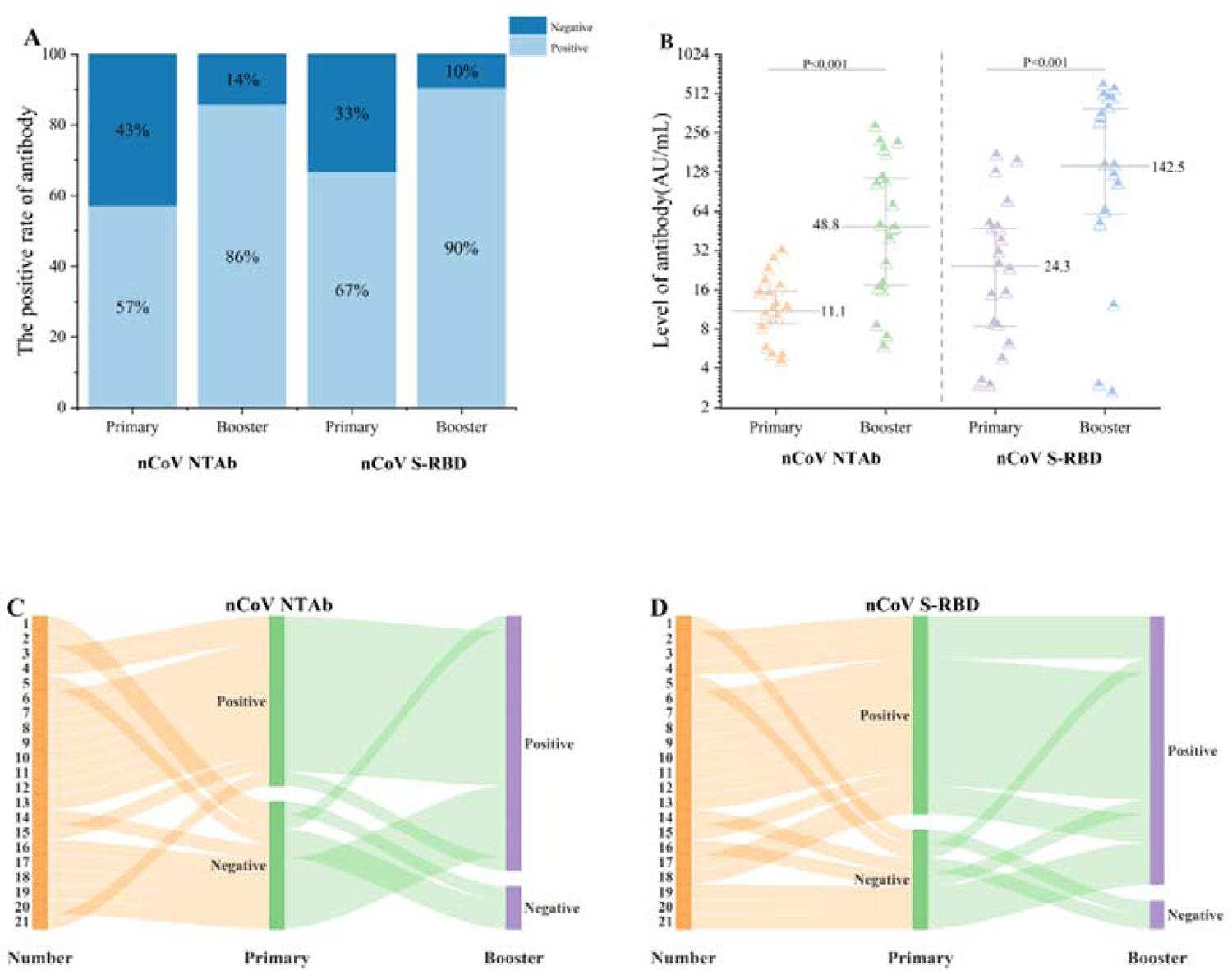
The dynamic changes of antibody in 21 patients performing twice antibody tests. Figure A revealed the positive rate of nCoV NTAb and nCoV S-RBD in Primary and Booster groups. Figure B revealed the level of nCoV NTAb and nCoV NTAb in Primary and Booster groups. Figure C revealed the dynamic change of nCoV NTAb after primary immunity and booster immunity. Figure D revealed the dynamic change of nCoV S-RBD after Primary immunity and booster immunity. Antibody level over 10.0 AU/mL was thought positive, while antibody level less than 10.0 AU/mL was thought negative. nCoV NTAb, novel coronavirus neutralizing antibody; nCoV S-RBD, novel coronavirus spike receptor-binding domain antibody.

**Table 4.** The dynamic changes of antibody in 21 patients performing twice antibody tests.

|  | Primary | Booster | P-Value |
| --- | --- | --- | --- |
| **nCoV NTAbs** |  |  |  |
| The positive rate | 12(57.1) | 18(85.7) | 0.085 |
| Antibody level | 11.1(8.4-16.0) | 48.8(16.8-143.4) | <0.001 |
| **nCoV S-RBD** |  |  |  |
| The positive rate | 14(66.7) | 19(90.5) | 0.133 |
| Antibody level | 24.3(7.2-48.9) | 142.5(55.8-424.9) | <0.001 |

Data was presented as median (interquartile range) or frequencies(%). Antibody level above 10.0 AU/mL was regarded as positive, while antibody level below 10.0 AU/mL was regarded as negative. nCoV NTAbs, novel coronavirus neutralizing antibody; nCoV S-RBD, novel coronavirus spike receptor-binding domain antibody.

After primary immunity, 9/21 cases were negative for nCoV NTAb, 7/21 were negative for nCoV S-RBD, and 6/21 patients were negative for both nCoV NTAb and nCoV S-RBD. After primary immunity, the median age of nCoV NTAb-negative patients was 57.0 (IQR: 46.5-66.0) years higher than 53.5 (IQR: 48.8-67.8) years in nCoV NTAb-positive patients, and the proportion of cirrhosis in nCoV NTAb-negative patients was 88.9%, which was higher than 50.0% in nCoV NTAb-positive patients (Table 5). After primary immunity, the median age of nCoV S-RBD-negative patients was 57.0 (IQR: 48.0–74.0) years higher than 53.5 (IQR: 47.8-67.3) years of nCoV S-RBD-positive patients, and the proportion of cirrhosis in nCoV S-RBD-negative patients was 85.7% higher than 57.1% in nCoV S-RBD-positive patients (Table 6).

**Table 5.**
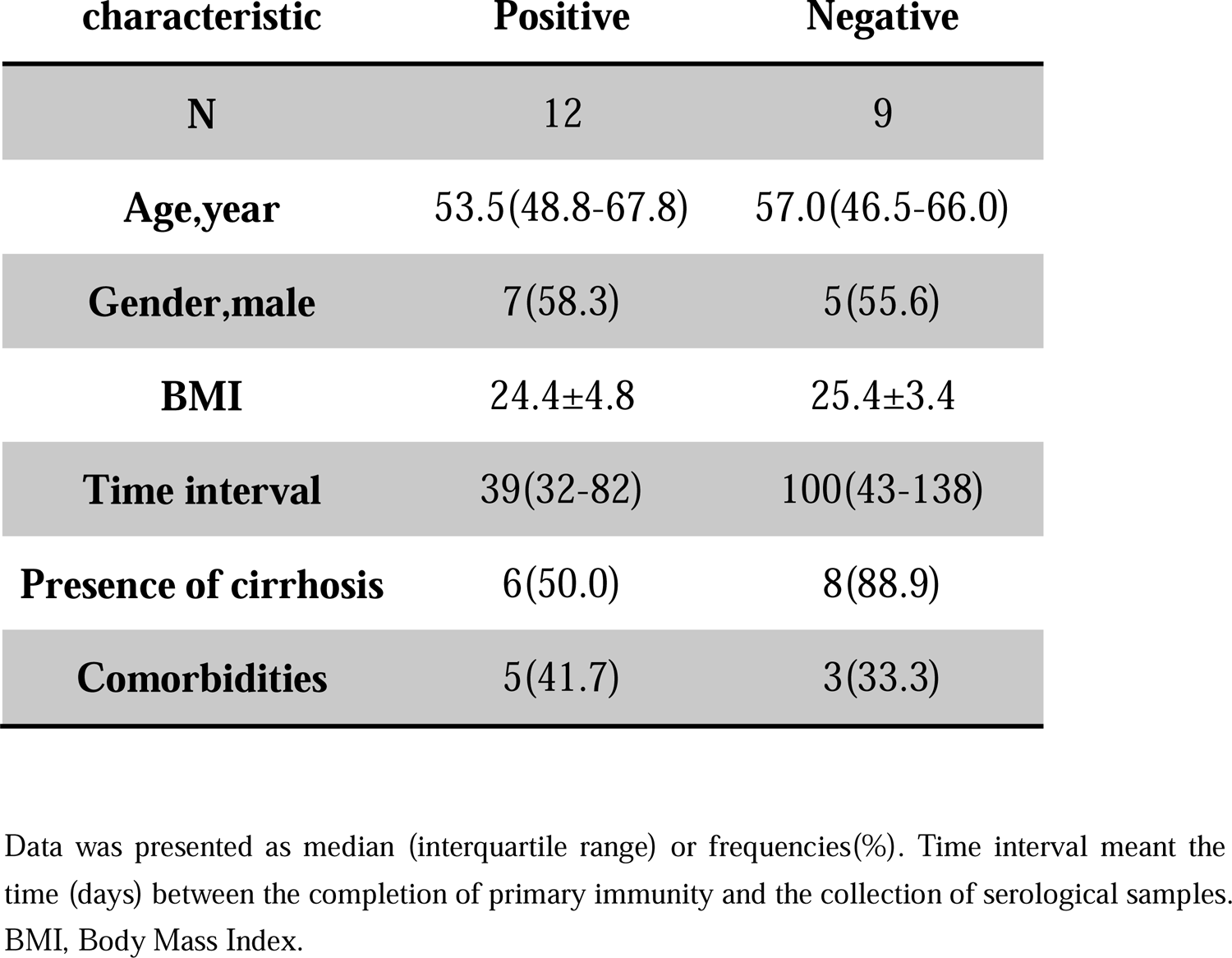
Demographic and clinical characteristics of nCoV NTAb positive and negative after primary immunity in 21 patients.

| **characteristic** | **Positive** | **Negative** |
| --- | --- | --- |
| **N** | 12 | 9 |
| **Age,year** | 53.5(48.8-67.8) | 57.0(46.5-66.0) |
| **Gender,male** | 7(58.3) | 5(55.6) |
| **BMI** | 24.4±4.8 | 25.4±3.4 |
| **Time interval** | 39(32-82) | 100(43-138) |
| **Presence of cirrhosis** | 6(50.0) | 8(88.9) |
| **Comorbidities** | 5(41.7) | 3(33.3) |

Data was presented as median (interquartile range) or frequencies(%). Time interval meant the time (days) between the completion of primary immunity and the collection of serological samples. BMI, Body Mass Index.

**Table 6.** Demographic and clinical characteristics of nCoV S-RBD positive and negative after primary immunity in 21 patients.

| **characteristic** | **Positive** | **Negative** |
| --- | --- | --- |
| **N** | 14 | 7 |
| **Age,year** | 53.5(47.8-67.3) | 57.0(48.0-74.0) |
| **Gender,male** | 8(57.1) | 4(57.1) |
| **BMI** | 24.9±4.7 | 24.6±3.3 |
| **Time interval** | 39(32-90) | 100(56-136) |
| **Presence of cirrhosis** | 8(57.1) | 6(85.7) |
| **Comorbidities** | 6(42.9) | 2(28.6) |

Data was presented as median (interquartile range) or frequencies(%). Time interval meant the time (days) between the completion of primary immunity and the collection of serological samples. BMI, Body Mass Index.

### 2.3 Safety assessment

After primary immunity, 46/203 (22.7%) participants happened the side effects. Local side effects occurred as follows: ache at the injection site (17, 8.4%), pruritus (2, 1.0%) and swelling (1, 0.5%). The most frequent systemic side effect was fatigue (15, 7.4%), followed by nausea (5, 2.5%), fever (4, 2.0%), dizziness (4, 2.0%), myalgia (4, 2.0%), decreased appetite (3, 1.5%), arthralgia (2, 1.0%), and cough (1, 0.5%). Local and systemic adverse reactions after primary immunity were slight and resolved over time.

After booster immunity, 29/113 (25.7%) participants reported the side effects. Local side effects occurred as follows: pain at the injection site (16, 14.2%), pruritus (1, 0.9%). The occurrence of systemic adverse reactions was as follows: fatigue (5, 4.4%), nausea (4, 3.5%), dizziness (3, 2.7%), decreased appetite (2, 1.8%), fever (1, 0.9%), myalgia (1, 0.9%), arthralgia (1, 0.9%), and cough (1, 0.9%). Adverse reactions after booster immunity were slight and self-limiting.

## 3. Discussion

The epidemic of COVID-19 has threatened public health; it is considered that vaccines have turned a criticial defence step. A prior investigation discovered that the effectiveness of SARS-CoV-2 vaccination was 95% in preventing COVID-19[13]. The positive rate of nCoV S-RBD was 99.6% in healthy people [14]. In another study, two doses of vaccines effectuated a serological conversion of antibodies up to 100% in healthy individuals[15].

Patients with comorbidities have a high morbidity of COVID-19 and are prone to develop severe illness and death[16]. The serological conversion rates of nCoV S-RBD in patients with HIV, hematopoietic stem cell transplantation, primary immunodeficiency diseases, chronic lymphocytic leukemia, and solid organ transplantation were 98.7%, 84.7%, 73.3%, 63.3%, and 43.4%, respectively, while that of the healthy control group was 100.0%[17]. After SARS-CoV-2 vaccination, the inhibitory level of nCoV NTAb in ovarian cancer patients was 83.6%, which was substantially lower than that of the healthy control group (92.9%)[18]. In this study, the positive serological rates of nCoV NTAb and nCoV S-RBD were 60.1% and 70.0% in CLD after primary immunity. Compared to a previous research[19], the positive rate of nCoV NTAb in this investigation was lower, which might be due to older age [54.0 (IQR: 42.0-62.0) years *vs*. 47.0 (IQR: 38.0–56.0) years], a larger percentage of cirrhosis (47.3 *vs*. 35.0), and a higher incidence of comorbidity (38.4 *vs*. 12.8).

Nonetheless, the effectiveness of SARS-CoV-2 vaccines diminished over time. The two doses of homologous ChAdOx1 vaccines were 68% effective in preventing COVID-19 at 15–30 days after administration, but no efficacy was detected at 121 days after administration[20]. Three doses of ZF2001 vaccines in preventing COVID-19 were 75.7% effective in healthy individals in 6 months after administration[21]. At six months after a booster dose, the positive serological rate of nCoV NTAb was >50%[22]. The third dose of the BNT162b2 vaccination increased the positive rate of nCoV S-RBD in patients undergoing liver transplantation from 56% to 98%[23]. After a booster dose, the level of nCoV NTAb in solid tumor patients was improved by three times compared to two doses but remained lower than the healthy control group[24]. Several investigations have revealed the humoral immune reaction of patients with CLD after primary immunity with SARS-CoV-2 vaccines. However, data regarding booster immunity on the humoral immune reaction in patients with CLD are lacking. In our research, the positive rate of nCoV NTAb was increased by 27.5%, and the antibody level was increased by 2.7 times after booster immunity. The positive rate of nCoV S-RBD was increased by 21.2%, and the antibody level was increased by 5.1-fold after booster immunity. In conclusion, after booster immunity, the humoral immune response in patients with CLD could be improved after primary immunity.

A total of 21 patients underwent antibody detection after both primary and booster immunities to further confirm the dynamic change in antibodies after booster immunity. Subsequently, the positive rate of nCoV NTAb increased from 57.1% to 85.7%, and the positive rate of nCoV S-RBD increased from 66.7% to 90.5%. The median level of nCoV NTAb increased from 11.1 AU/mL to 48.8 AU/mL after booster immunity. The median level of nCoV S-RBD was increased from 24.3 AU/mL to 142.5 AU/mL. Compared to the antibody level of primary immunity, the median level of nCoV NTAb and nCoV S-RBD was increased by 4.4- and 5.9-fold, respectively. Consistent with the literature[25], patients with CLD improve the humoral immune response after completing booster immunity, and the antibody level of patients with autoimmune hepatitis negative for nCoV S-RBD was increased by 148-fold[25]. Booster immunity prompted an immune response in liver transplant patients negative for nCoV S-RBD[23]. 9/21 patients were negative for nCoV NTAb after primary immunity, while 6 cases were transformed positive after booster immunity, and the positive conversion rate of nCoV NTAb was 66.7%. A total of 7 patients were negative for nCoV S-RBD after primary immunity, while 5 cases were transformed positive after booster immunity; the positive conversion rate of nCoV S-RBD was 71.4%. The positive conversion rate among patients with primary immunity failure can be improved after booster immunity, such that patients with primary immunity failure can obtain immunity to COVID-19.

In this study, we executed the subgroup analysis to contrast the immune reaction in patients with non-cirrhosis and cirrhosis in both Primary and Booster groups. Consistent with previous studies[6, 19], the positive rate and level of nCoV NTAb and nCoV S-RBD had no significant difference between non-cirrhosis and cirrhosis patients in Primary. However, the positive rate of nCoV NTAb and nCoV S-RBD without cirrhosis was higher than that in patients with cirrhosis in Booster. Patients with cirrhosis have congenital and adaptive immune dysfunction[26] and B cell phenotypic and functional defects[27]. The number of total T lymphocytes decreases in patients with cirrhosis, while CD4+ and CD8+ T cells expressing apoptosis markers are significantly increased[28]. These explain the unsatisfactory immune response of patients with cirrhosis after vaccines.

In the current review, we examined the data of 21 patients whose antibodies were negative after primary and booster immunity, respectively. Due to the small samples, statistical analysis was not conducted. We found that patients with negative SARS-CoV-2 antibodies after primary immunity were older and had a high proportion of cirrhosis than those with positive antibodies. After booster immunity, 3 were negative for SARS-CoV-2 antibody, 1 was negative only for nCoV NTAb, and 2 were negative for both nCoV NTAb and nCoV S-RBD. First, 3 cases were cirrhotic and older. In patients with only nCoV NTAb negative, the level of nCoV S-RBD was 11.9 AU/mL. The patient was 74-years-old and had a BMI of 31.6, so the effect of SARS-CoV-2 vaccine was not optimal after booster immunity. Previous studies reported that age and BMI were related to the antibody level [6, 23, 29], suggesting that elderly with underlying diseases, such as cirrhosis and obesity, should receive booster vaccinations regularly to obtain sustained immunity against COVID.

Consistent with the results of current studies[6, 19], most of the local and systemic side effects in patients with CLD after primary immunity were slight and could be alleviated in this study. The most frequent local and systemic side effect after primary immunity were pain at the injection site and fatigue, respectively. Reportedly, the side effects of healthy individuals are slight after booster immunity[21, 22]. In this study, we found that the side effects of patients with CLD after booster immunity were mostly mild and self-limited. The most frequent local and systemic adverse reaction after booster immunity were pain at the injection site and fatigue, respectively.

The level of nCoV NTAb predicts vaccine protection[30]. Regarding the gold standard for the detection of nCoV NTAb[31, 32], plaque reduction neutralization test (PRNT) involves live virus, and the cell culture and low detection throughput require a high laboratory level and complex procedure. In the present study, competitive combined chemiluminescence immunoassay (CLIA) was used, which has the characteristics of high automation and throughput[33]. A major target of the SARS-CoV-2 vaccine is the S protein, especially the RBD region on the S protein[34]. A strong correlation was established between nCoV NTAb and nCoV S-RBD, and it has been confirmed that the detection of nCoV SRBD antibodies is highly-sensitive and specific[35]. Therefore, two types of antibodies, nCoV NTAb and nCoV S-RBD, were determined in this study.

Nevertheless, there are a few drawbacks in current study. It was a single-center study, and the samples were from the same region. Thus, samples from other regions should be enrolled in the future to make the research results universal. During this study, only the humoral immunity status of patients with CLD who completed SARS-CoV-2 vaccination was estimated, and the cellular immunity status of patients was not evaluated, which is crucial to understand the immunogenicity of SARS-CoV-2 vaccine. Only patients with CLD were included, and healthy individuals were not included as the control group, which did not allow the comparison of the differences in the immunogenicity of SARS-CoV-2 vaccine between patients with CLD and healthy individuals. In the future, we would need to include additional specimens with populations from other areas to confirm our conclusions further.

To date, some countries have started to vaccinate high-risk groups with the fourth dose[36]; this study provided data support for the formulation of vaccination strategies. In conclusion, patients with CLD improved the humoral immune response after completing primary and booster immunity of SARS-CoV-2 vaccines, while booster immunity further improved the positive rate and antibody level of patients with CLD. Finally, the positive conversion rate among patients with primary immunity failure could be improved after booster immunity.

## Data Availability

All data produced in the present study are available upon reasonable request to the authors
All data produced in the present work are contained in the manuscript
All data produced are available online

